# Prioritisation of potential anti-SARS-CoV-2 drug repurposing opportunities based on ability to achieve adequate plasma and target site concentrations derived from their established human pharmacokinetics

**DOI:** 10.1101/2020.04.16.20068379

**Authors:** Usman Arshad, Henry Pertinez, Helen Box, Lee Tatham, Rajith KR Rajoli, Paul Curley, Megan Neary, Joanne Sharp, Neill J Liptrott, Anthony Valentijn, Christopher David, Steve P Rannard, Paul O’Neill, Ghaith Aljayyoussi, Shaun Pennington, Stephen A Ward, David J Back, Saye H Khoo, Patrick G Bray, Giancarlo Biagini, Andrew Owen

**Affiliations:** Department of Molecular and Clinical Pharmacology, University of Liverpool, Liverpool, L7 3NY, UK; Department of Chemistry, University of Liverpool, Liverpool, L69 3BX, UK; Centre for Drugs and Diagnostics. Liverpool School of Tropical Medicine, Liverpool L3 5QA, UK; Pat Bray Electrical, 260D Orrell Road, Orrell, Wigan, WN5 8QZ, UK

## Abstract

There is a rapidly expanding literature on the *in vitro* antiviral activity of drugs that may be repurposed for therapy or chemoprophylaxis against SARS-CoV-2. However, this has not been accompanied by a comprehensive evaluation of the ability of these drugs to achieve target plasma and lung concentrations following approved dosing in humans. Moreover, most publications have focussed on 50% maximum effective concentrations (EC_50_), which may be an insufficiently robust indicator of antiviral activity because of marked differences in the slope of the concentration-response curve between drugs. Accordingly, *in vitro* anti-SARS-CoV-2 activity data was digitised from all available publications up to 13^th^ April 2020 and used to recalculate an EC_90_ value for each drug. EC_90_ values were then expressed as a ratio to the achievable maximum plasma concentrations (Cmax) reported for each drug after administration of the approved dose to humans (Cmax/EC_90_ ratio). Only 14 of the 56 analysed drugs achieved a Cmax/EC_90_ ratio above 1 meaning that plasma Cmax concentrations exceeded those necessary to inhibit 90% of SARS-CoV-2 replication. A more in-depth assessment of the putative agents tested demonstrated that only nitazoxanide, nelfinavir, tipranavir (boosted with ritonavir) and sulfadoxine achieved plasma concentrations above their reported anti-SARS-CoV-2 activity across their entire approved dosing interval at their approved human dose. For all drugs reported, the unbound lung to plasma tissue partition coefficient (K_p_U_lung_) was also simulated and used along with reported Cmax and fraction unbound in plasma to derive a lung Cmax/EC_50_ as a better indicator of potential human efficacy (lung Cmax/EC_90_ ratio was also calculable for a limited number of drugs). Using this parameter hydroxychloroquine, chloroquine, mefloquine, atazanavir (boosted with ritonavir), tipranavir (boosted with ritonavir), ivermectin, azithromycin and lopinavir (boosted with ritonavir) were all predicted to achieve lung concentrations over 10-fold higher than their reported EC_50_. This analysis was not possible for nelfinavir because insufficient data were available to calculate K_p_U_lung_ but nitozoxanide and sulfadoxine were also predicted to exceed their reported EC_50_ by 3.1- and 1.5-fold in lung, respectively. The antiviral activity data reported to date have been acquired under different laboratory conditions across multiple groups, applying variable levels of stringency. However, this analysis may be used to select potential candidates for further clinical testing, while deprioritising compounds which are unlikely to attain target concentrations for antiviral activity. Future studies should focus on EC_90_ values and discuss findings in the context of achievable exposures in humans, especially within target compartments such as the lung, in order to maximise the potential for success of proposed human clinical trials.

## Introduction

Coronavirus 2019 (COVID-19) is a respiratory disease caused by severe acute respiratory syndrome coronavirus 2 (SARS-CoV-2) infection. Fever, a persistent cough and respiratory symptoms are common, with some patients reporting vomiting, nausea, abdominal pains and diarrhoea [1]. To date, no specific treatment is available, and this has resulted in significant morbidity and mortality globally. According to the International Clinical Trials Registry Platform search portal, 927 clinical trials for COVID-19 have been registered [2]. This rapidly expanding pandemic warrants the urgent development of strategies, particularly to protect people at high risk of infection. Repurposing currently available drugs that have been utilised clinically with a known safety profile, is the quickest way to address this serious unmet clinical need. Antiviral drugs are urgently required for treatment of patients with mild/moderate disease to prevent the worsening of symptoms and reduce the burden upon healthcare systems. However, a different approach is likely to be needed for patients that are already in a critical state, due to the immune dysregulation which is so apparent in severe cases [3].

Previous investigations have shown that the entry by SARS-CoV occurs via the angiotensin converting enzyme 2 (ACE2) receptor [4]. A study on normal lung tissue showed that 83% of ACE2-expressing cells were alveolar epithelial type II cells [5], highlighting the lungs as the primary target organ that facilitate viral invasion and replication. Furthermore, the ACE2 receptor is also highly expressed in gastrointestinal epithelial cells, with SARS-CoV-2 RNA observed to be present in stool specimens of patients during infection [1, 6]. A recent retrospective analysis of 85 patients with laboratory confirmed COVID-19 also indicated that SARS-CoV-2 infects human kidney tubules and induces acute tubular damage in some patients [7]. Furthermore, 2–11% of patients with COVID-19 exhibit liver comorbidities [8]. Of note is an observation of SARS and Middle East respiratory syndrome (MERS) having a tropism to the gastrointestinal tract [9] and causing liver impairments in addition to respiratory disease. The genomic similarity between SARS-CoV-2 and SARS-CoV (79.6% sequence identity) would imply that the current virus would act in a similar manner and be present within the body systemically [10-12]. Therefore, treatment options that provide therapeutic concentrations of drug(s) within the systemic circulation and other affected organs are likely to be required.

In the absence of a vaccine, antiviral drugs could also be deployed as chemoprophylaxis to protect against infection and would present an essential tool for protecting healthcare staff and other key workers, as well as household contacts of those already infected. For chemoprevention drugs will need to penetrate into the multiple sites where SARS-CoV-2 infection occurs, and do so in sufficient concentrations to inhibit viral replication [13]. This may include the mucous membranes present in the nasal cavity and throat, the ocular surface, tears and the upper respiratory tract/lungs [14, 15]. However, therapeutic concentrations may not be needed in the systemic circulation for chemoprophylaxis, but this is yet to be determined. Although difficult and scarcely studied, work in animals has shown that the size of the inoculum of other respiratory viruses such as influenza is associated with the severity of the resultant disease [16, 17]. Reports with SARS-CoV-2 indicate that higher viral loads are indicative of poorer prognosis and correlate with the severity of symptoms, with viral load in severe cases reported to be 60 times higher than that of mild cases [18, 19]. In light of this, even if a chemoprophylactic drug reduced inoculum size without completely blocking transmission, major benefits for morbidity and mortality may still be achievable.

Many ongoing global research efforts are focused on screening the activity of existing compounds *in vitro* in order to identify candidates to repurpose for SARS-CoV-2. However, current data have not yet been systematically analysed in the context of the plasma and target site exposures that are achievable after administration of the approved doses to humans. The purpose of this work was to evaluate the existing *in vitro* anti-SARS-CoV-2 data to determine and prioritise drugs capable of reaching antiviral concentrations within the blood plasma. An accepted physiologically-based pharmacokinetic (PBPK) equation was also used to predict the expected concentration in lung, in order to assess the potential of these drugs for therapy in this key disease site and the potential for chemoprevention.

## Methods

### Candidate Analysis

To identify compounds and their relevant potency and pharmacokinetic data, we performed a literature search on PubMed, Google Scholar, BioRxiv, MedRxiv, and ChemRxiv. The following search terms were used for *in vitro* activity data – (COVID-19 OR SARS-CoV-2) AND (EC50 OR IC50 OR antiviral). For pharmacokinetic data (Cmax OR pharmacokinetics) was used along with the drug name for drugs with reported anti-SARS-CoV-2 activity (up to April 13^th^ 2020). Further clinical pharmacokinetic data was obtained from the Food and Drug Administration (FDA), the European Medicines Agency (EMA) and through publications available online.

### Lung accumulation prediction

An indication of the degree to which candidate drugs are expected to accumulate in lung (a presumed site of primary efficacy and for prevention of SARS-CoV-2 infection) was provided by calculation of unbound lung to plasma tissue partition coefficient (K_p_U_lung_) according to the methodology of Rodgers and Rowland [20-22]. The formulae provided in these methods use the physicochemical properties of the drug (pKa, log P, classification as acid/base/neutral) and *in vitro* drug binding information (fraction unbound in plasma, blood to plasma ratio), in combination with tissue specific data (lipid content, volumes of intra/extracellular water etc.) to predict tissue K_p_U values. Predicted K_p_U_lung_ was converted to K_p_lung_ by multiplying by fraction unbound in plasma to allow estimation of lung exposure from *in vivo* measurements of plasma concentration. In the absence of observed tissue distribution data the Rodgers and Rowland method is an accepted means to provide initial estimates of tissue partitioning for PBPK modelling, but with known limits on potential accuracy (generally predicted K_p_U is within 2-3 fold of observed tissue K_p_U values).

### Data analysis and interpretation

Since in the majority of papers only an EC_50_ value was available, concentration-response data were digitized using the Web Plot Digitizer® software. Graphs were then re-plotted in SigmaPlot 14.0 (Systat Software, Inc.) and curves were fitted to confirm EC_50_ values and determine EC_90_ values. A Cmax/EC_50_ and Cmax/EC_90_ ratio was then calculated for each drug for which previous evidence of clinical use in humans and availability of human pharmacokinetic data were available. Lung K_p_U values were also used in combination with reported Cmax values to derive an estimate of lung exposure at Cmax for each drug. For a subset of molecules, the absence of available physicochemical or plasma protein binding parameters prohibited derivation of a K_p_U estimate. For the remaining drugs, a lung Cmax/EC_50_ and lung Cmax/EC_90_ was calculated. Published plasma concentration – time data for the most promising candidates were also digitized (were available) and replotted to visually represent human pharmacokinetics relative to the calculated EC_50_ and EC_90_ data.

## Results

### Identified papers and methods

We identified 14 key studies that detailed the antiviral activity of 72 compounds [23-37]. The majority of the *in vitro* SARS-CoV-2 infection experiments were performed in Vero E6 cells (ATCC 1586) maintained in either DMEM or MEM. Other studies utilised Vero-hSLAM cells, VeroE6 cells expressing TMPRSS2 and the CACO-2 cell line to cultivate the virus. The following SARS-CoV-2 strains were used across studies; WA-1 strain - BEI #NR-52281; Brazil/RJ-314/2020; C-Tan-nCoV Wuhan strain 01; Wuhan/WIV04/2019; USA-WA1/2020; nCoV-2019BetaCoV/Wuhan/WIV04/2019; BetaCoV/Hong Kong/VM20001061/2020; Australia/VIC01/2020; βCoV/KOR/KCDC03/2020 and BavPat1/2020. Cells across all studies were infected with the virus with a multiplicity of infection (MOI) of 0.002, 0.01, 0.0125, 0.02, 0.05 and 0.1. Drugs were added at concentrations varying between 0.01μM - 500μM. A summary of the differences in methodologies between studies reporting SARS-CoV-2 antiviral activity is presented in Table 1.

**Table 1.**
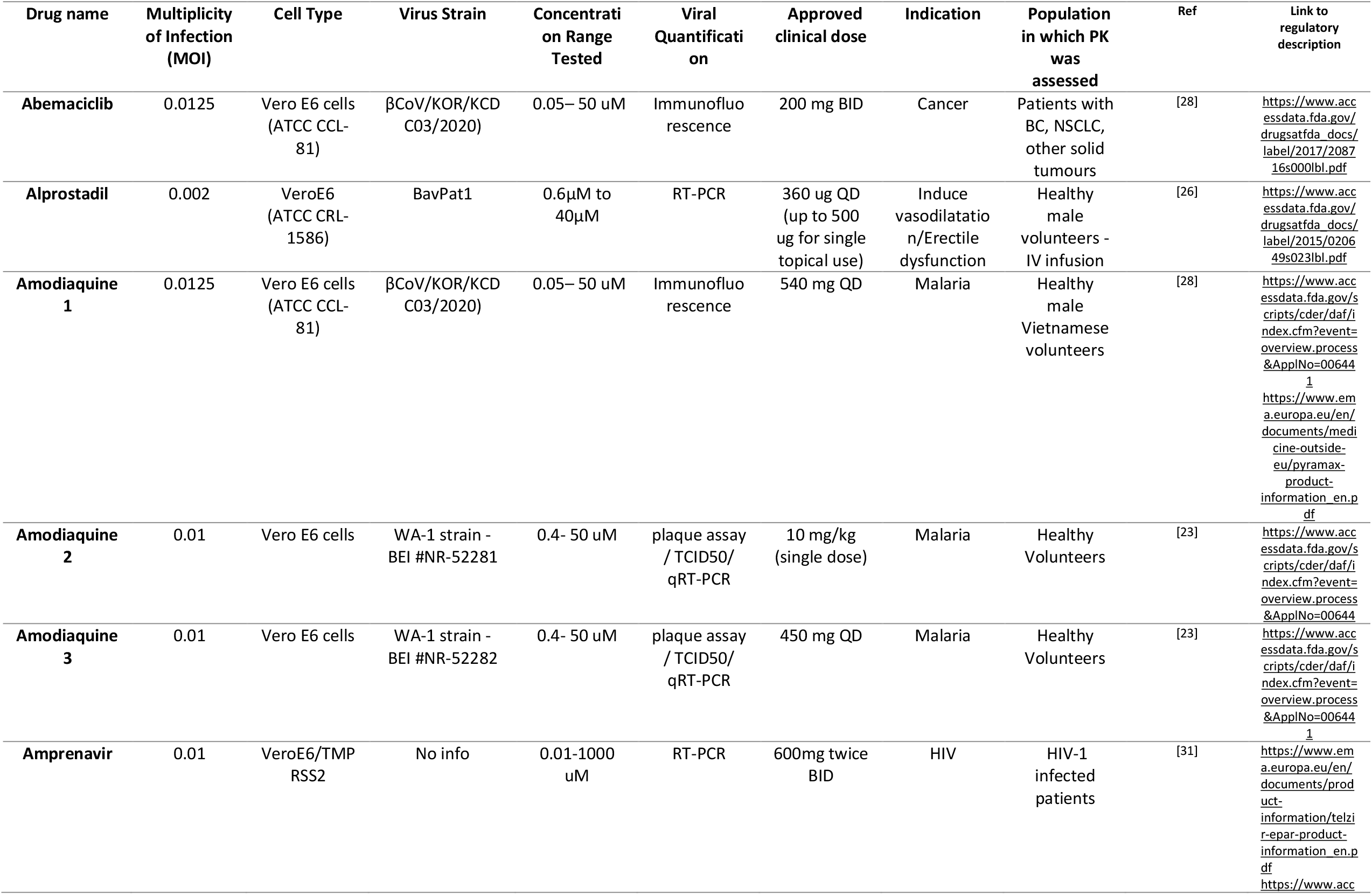
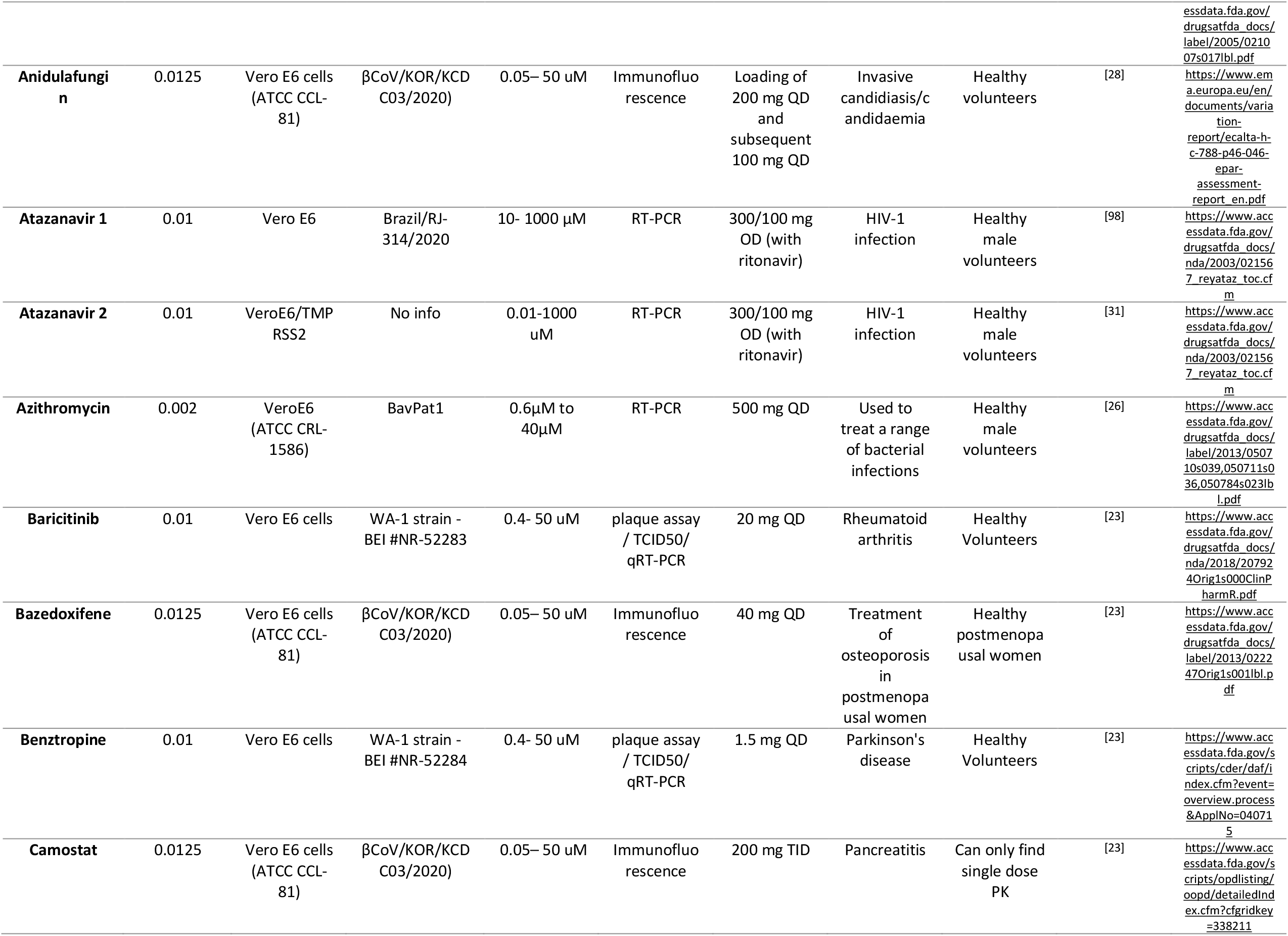
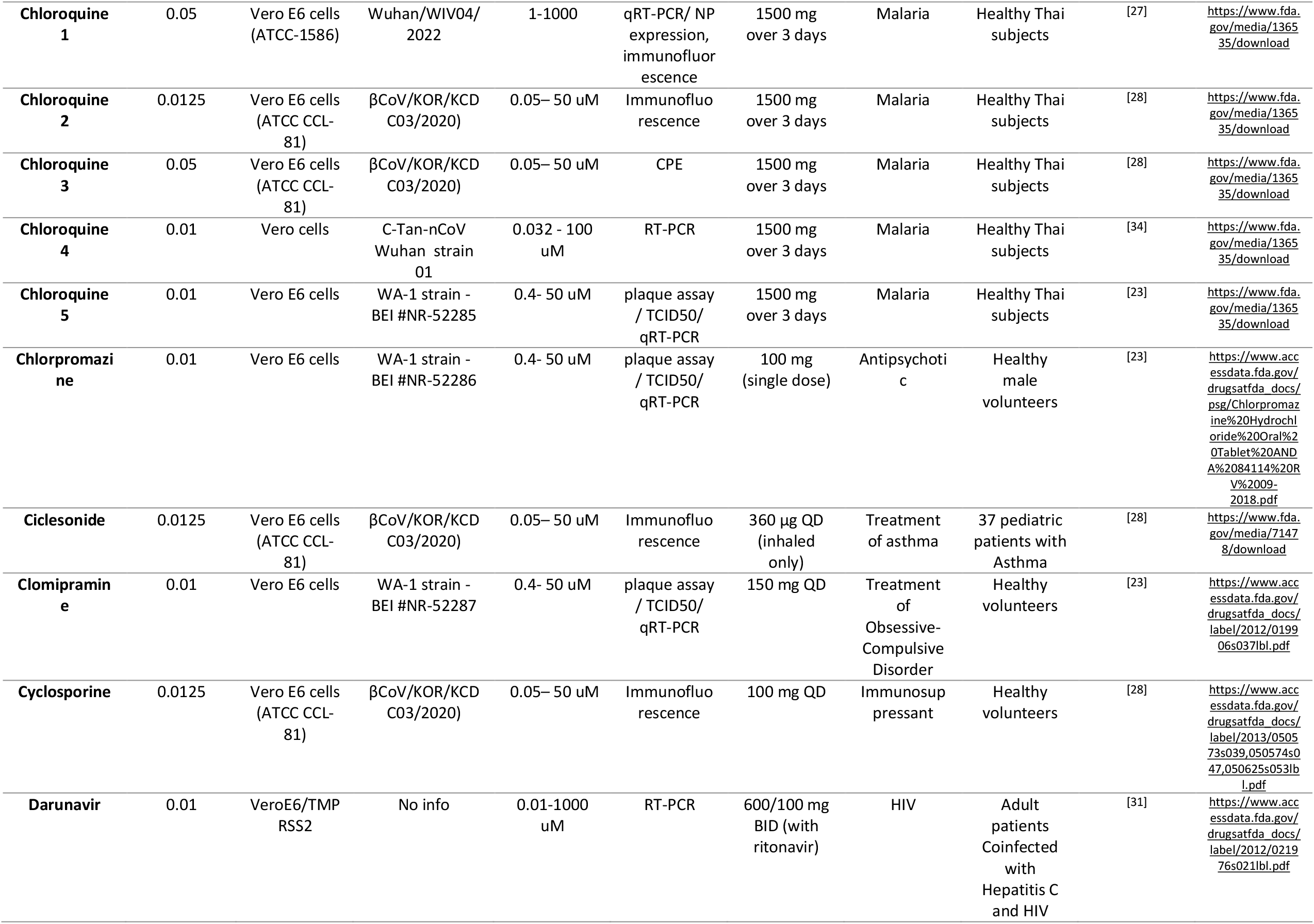
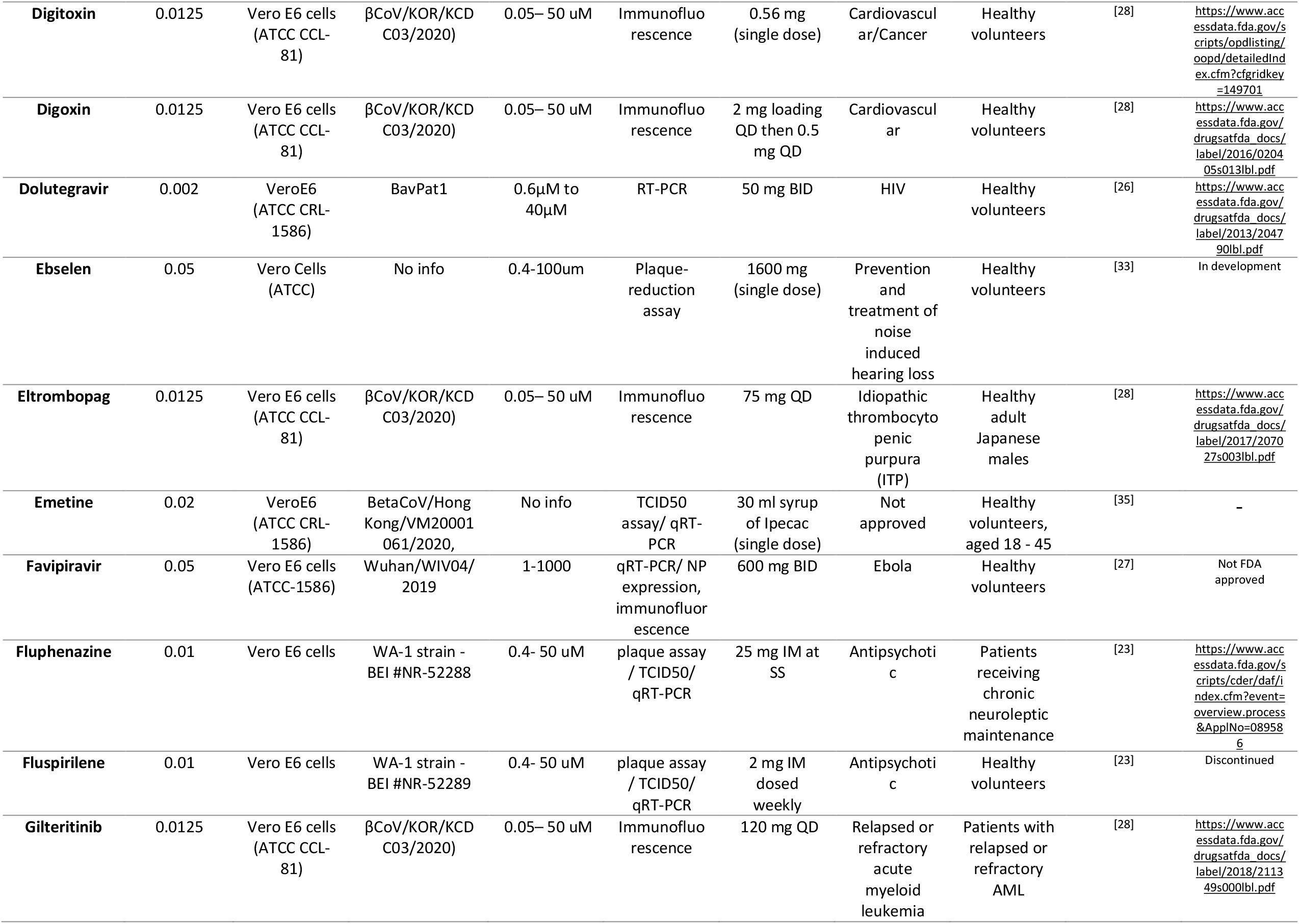
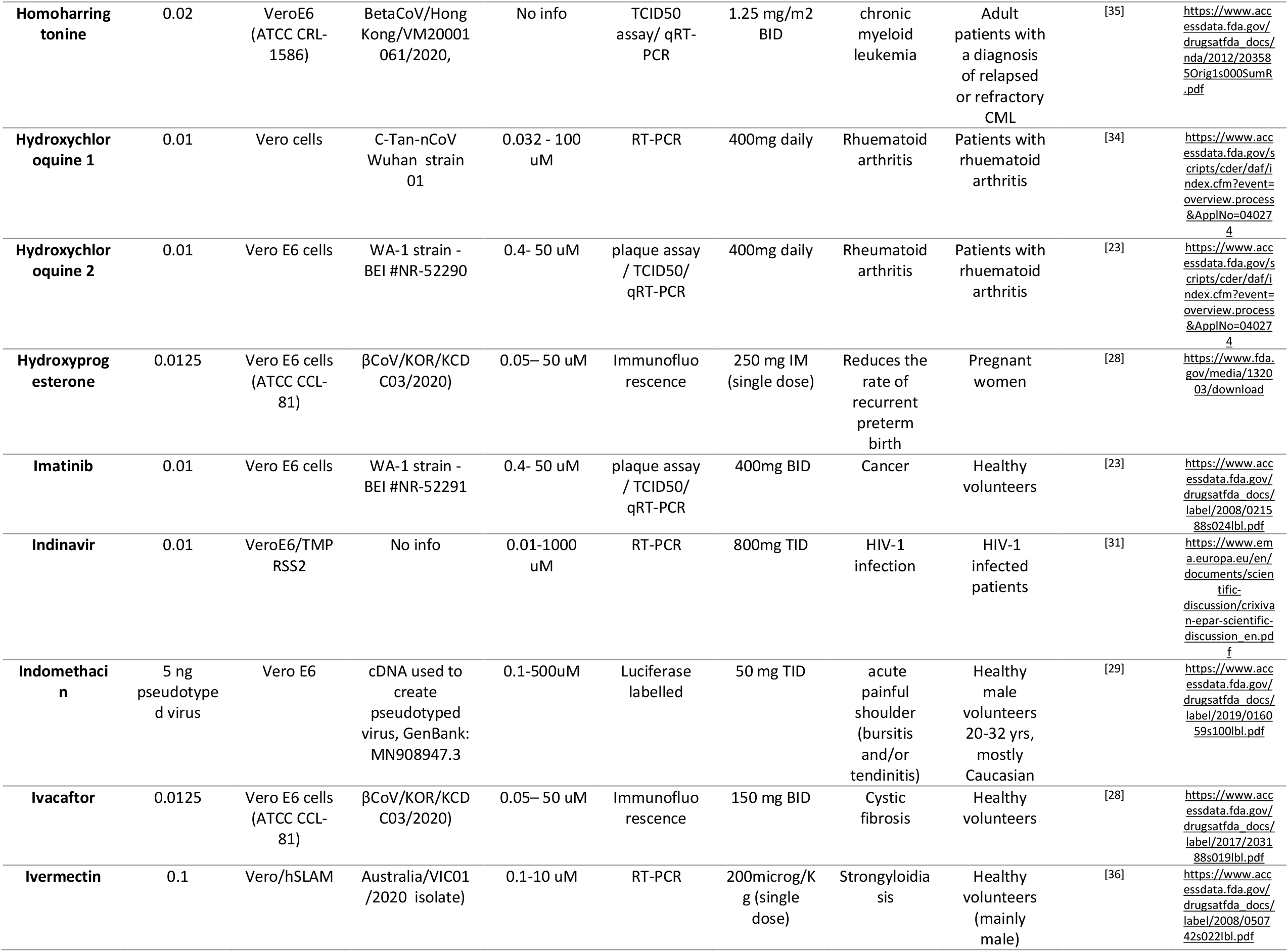
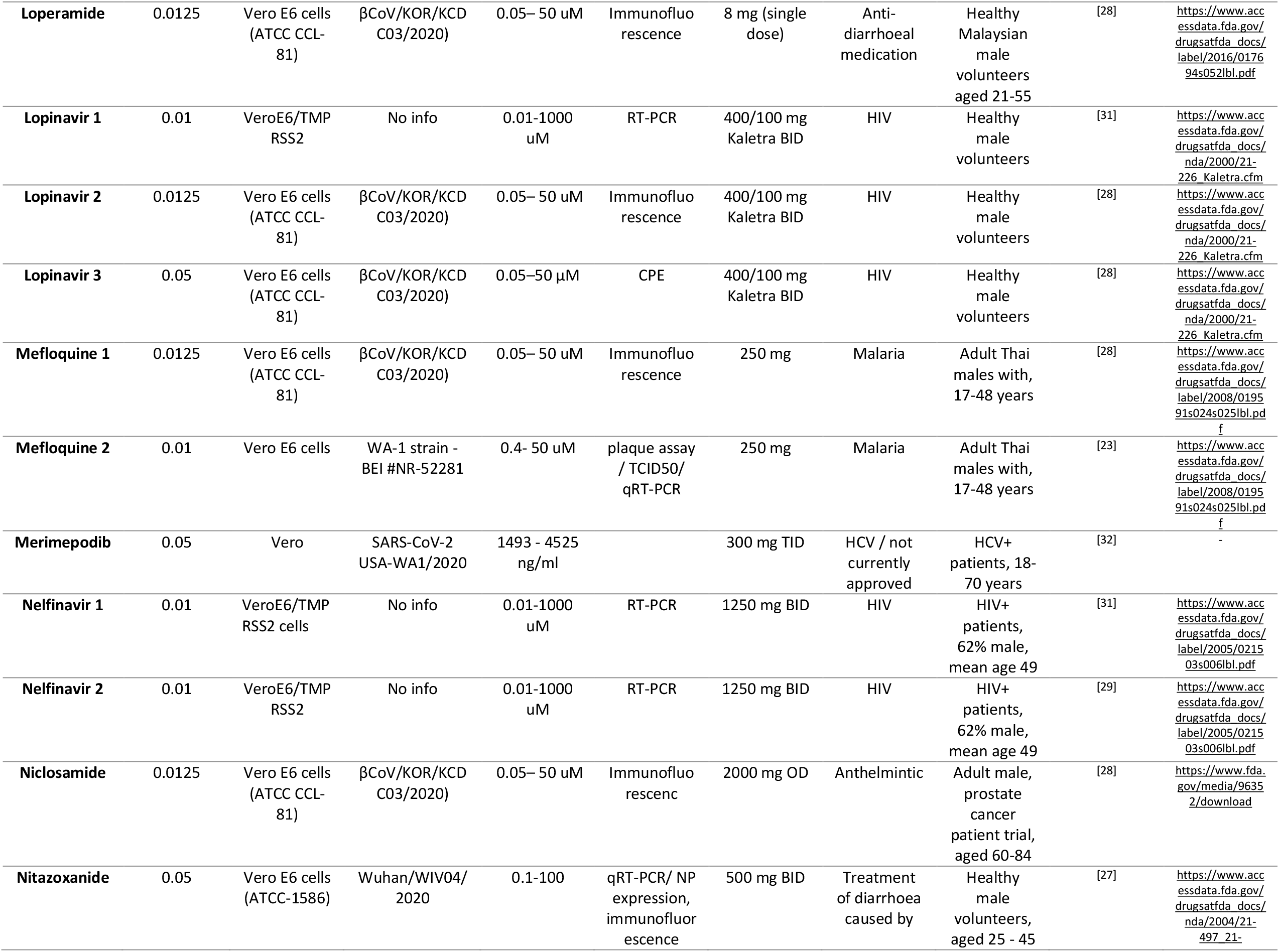
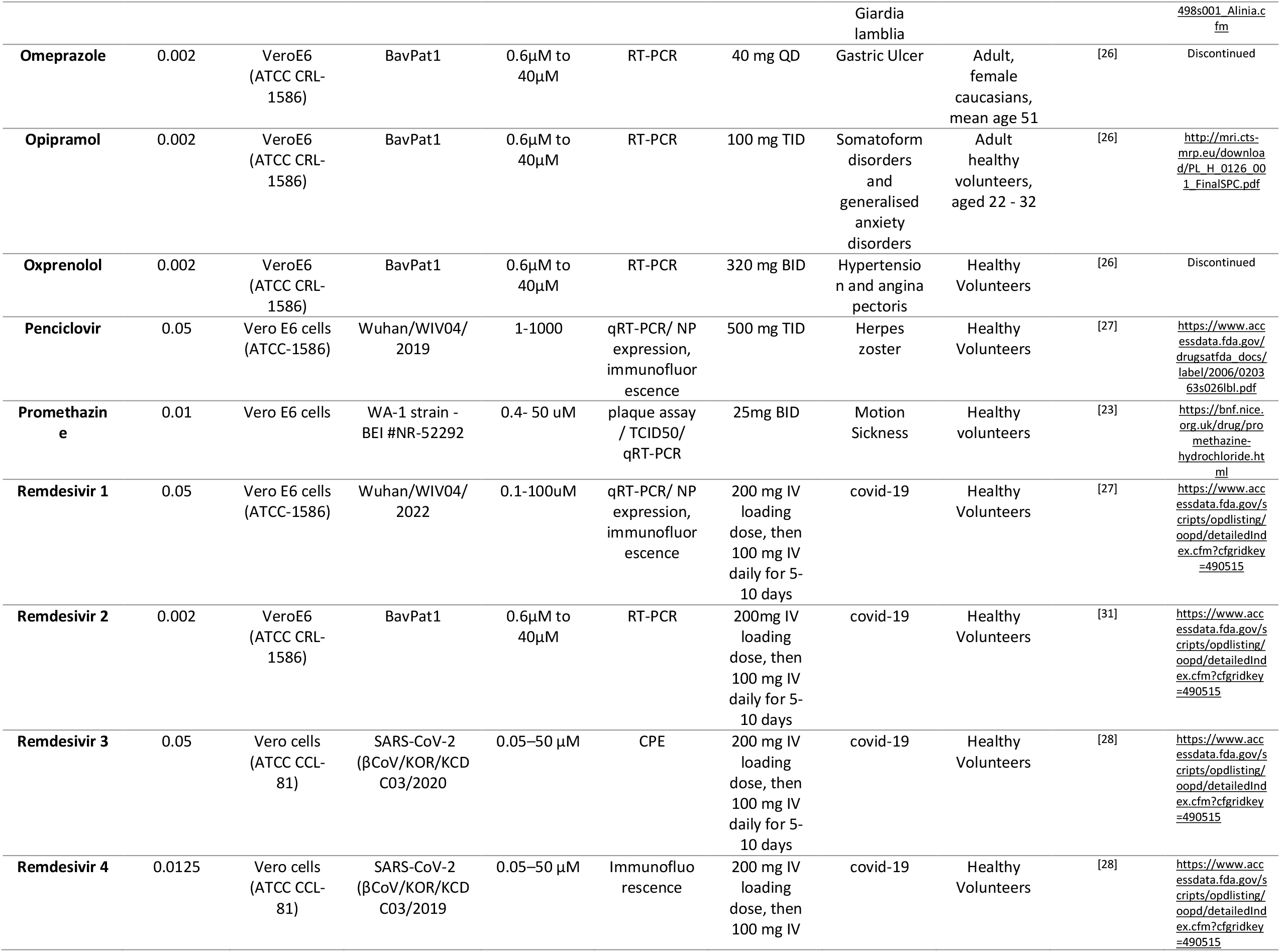
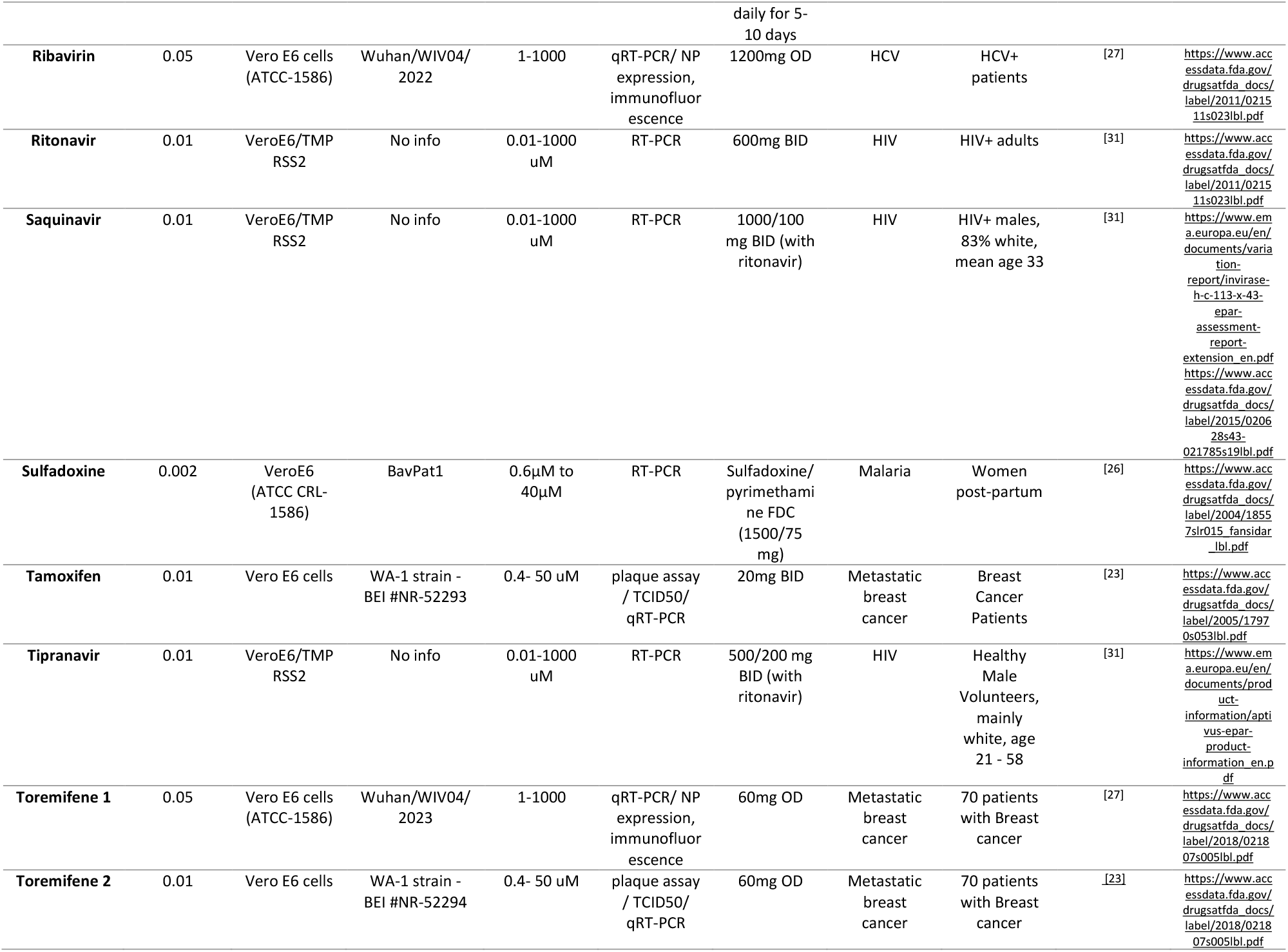
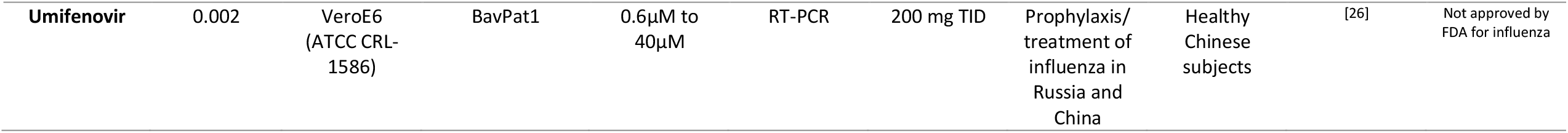
Drugs investigated for SARS-CoV-2 antiviral activity and experimental overview

**Table 2.**
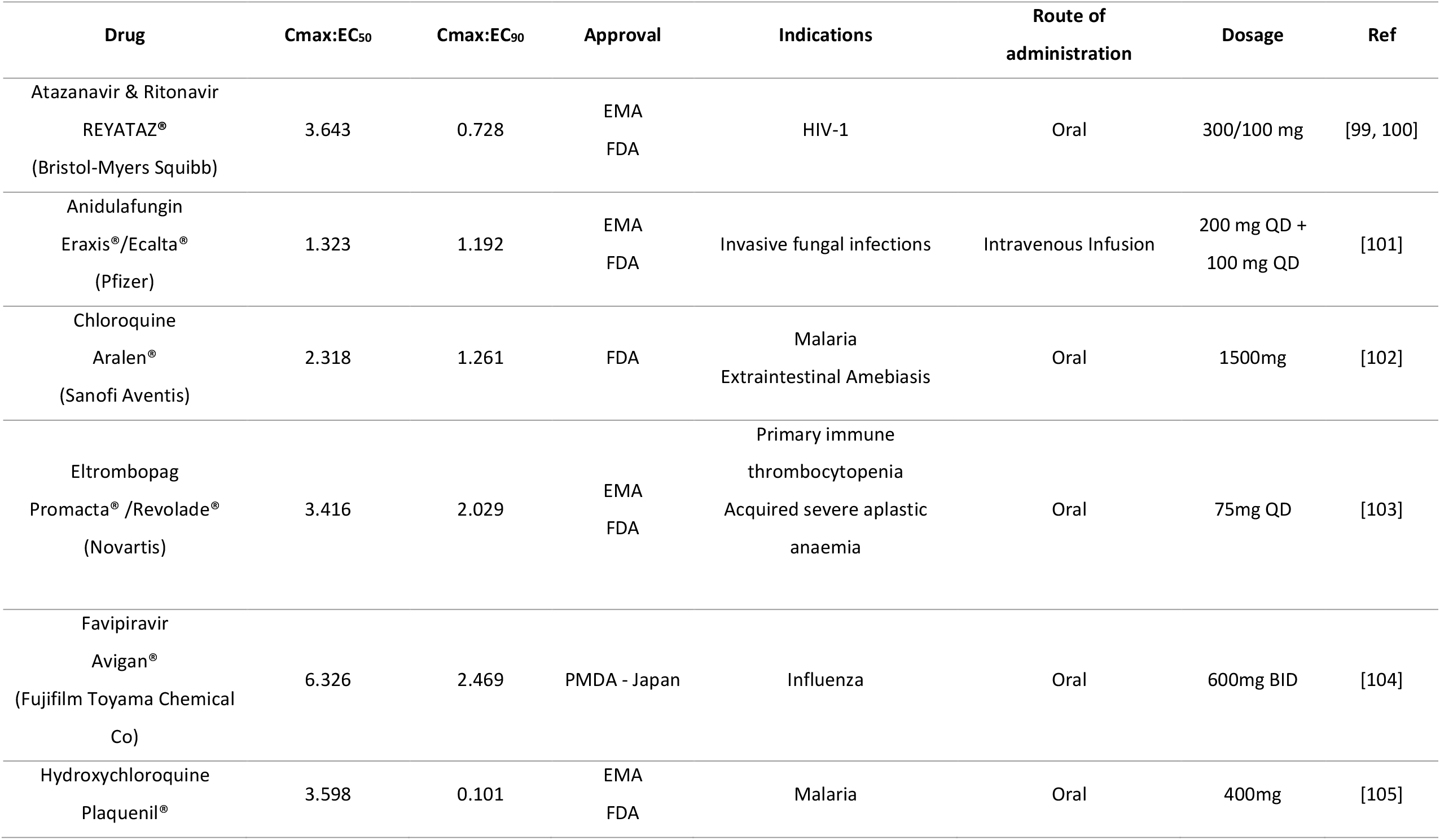
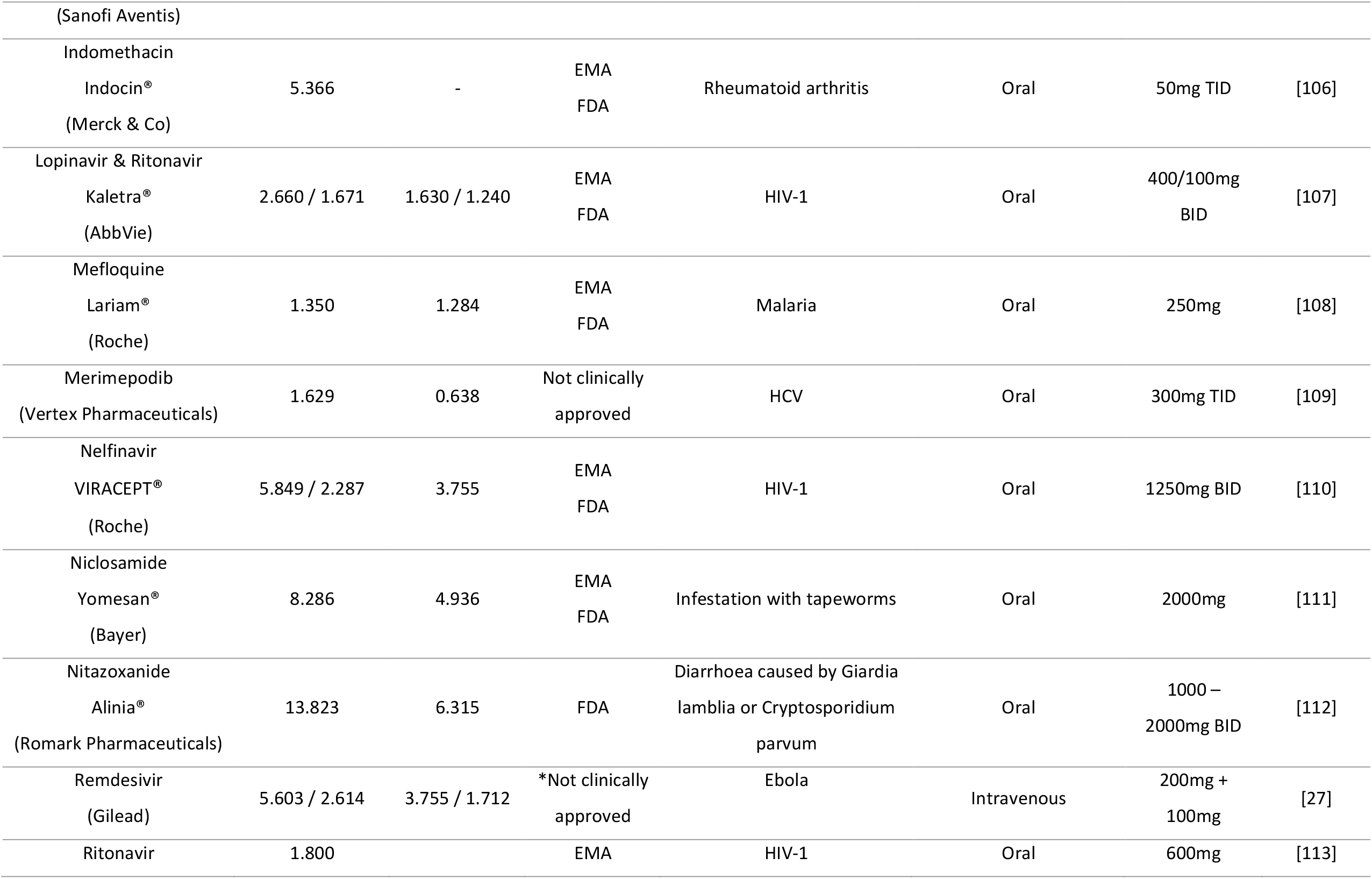
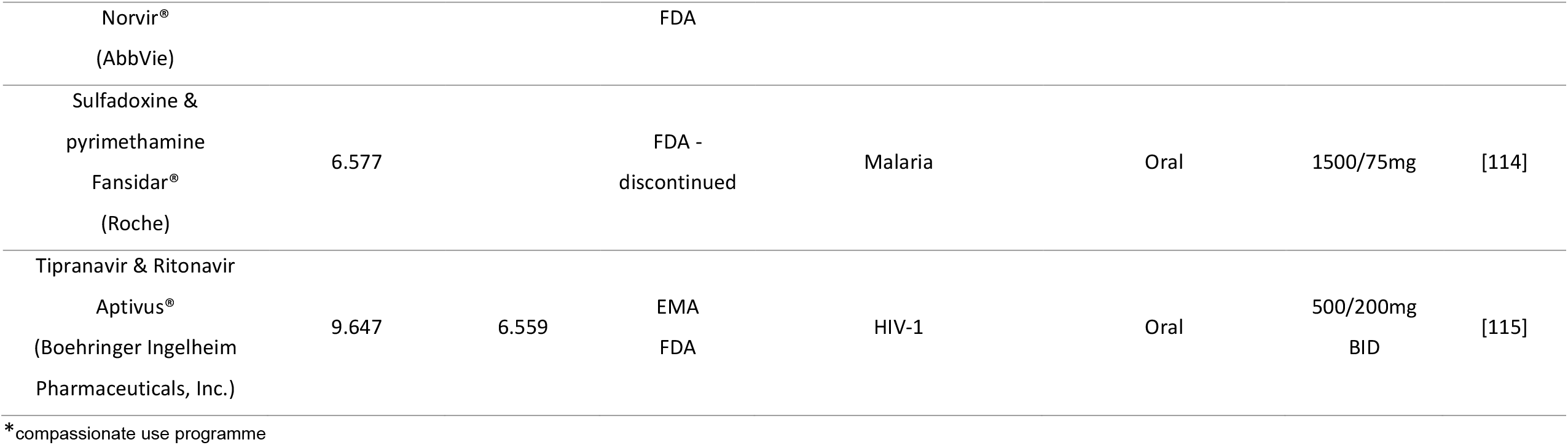
Summary of the top leads identified

### Identification of candidates achieving plasma concentrations expected to exert antiviral activity (Cmax / EC_50_ ratio)

Seventeen molecules had a reported Cmax value greater than at least one of the reported EC_50_ values against SARS-CoV-2 and these included nelfinavir, chloroquine, remdesivir, lopinavir (boosted with ritonavir), eltrombopag, hydroxychloroquine, atazanavir (boosted with ritonavir), indomethacin, favipiravir, sulfadoxine, nicolsamide, mefloquine, tipranavir (boosted with ritonavir), ritonavir, merimepodib, anidulafungin and nitazoxanide. However, it should be noted that for amodiaquine, atazanavir, chloroquine, hydroxychloroquine, lopinavir, mefloquine, nelfinavir, remdesevir and toremefine, more than one EC_50_ value had been reported across the available literature and these were not always in agreement (Figure 1A). Moreover, this variability in reported EC_50_ values sometimes resulted in Cmax / EC_50_ ratios giving a different estimation of the likely value of the molecule. Meaning that for the same drug, the Cmax / EC_50_ ratio could be above or below 1 (Figure 1B). For amodiaquine and toremifene all reported EC_50_ values were below their reported Cmax and only for nelfinavir was the reported Cmax expected to exceed both reported EC_50_ values. For atazanavir, chloroquine, hydroxychloroquine, lopinavir, mefloquine and remdesivir, some EC_50_ values were above the Cmax whereas other were below. This observation dramatically highlights the sensitivity of the current analysis to the reported antiviral activity data, and this should be taken into account when interpreting the data presented hereafter.

**Figure 1.**
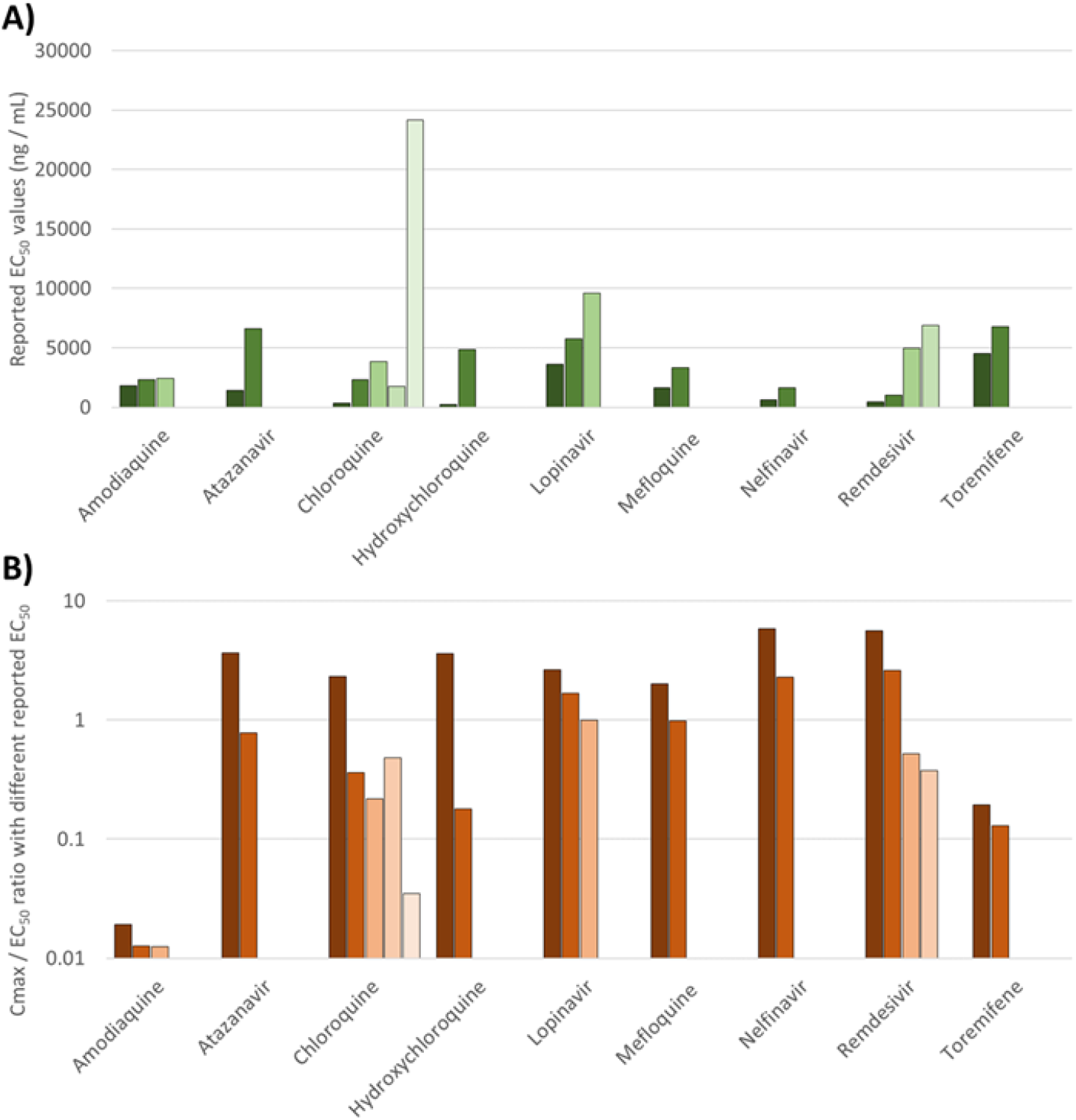
Assessment of the variation in reported EC_50_ values for SARS-CoV-2 across the drugs for which more than one value was available in the literature (A). The consequences of this variability in reported EC_50_ in terms of the Cmax / EC_50_ ratio is also provided (B). Amodiaquine and toremifene were estimated to exhibit sub-therapeutic pharmacokinetics irrespective of which EC_50_ value was used. Similarly, nelfinavir was estimated to have Cmax value higher than its EC_50_ irrespective of which EC_50_ was used in the analysis. For the other drugs, interpretation was highly dependent upon which reported EC_50_ was utilised and this underscores the caution that should be taken in interpreting the available data.

### Identification of candidates achieving plasma concentrations exceeding the SARS-CoV-2 EC_90_ (Cmax / EC_90_ ratio)

For 57 of the reported antiviral activities, data covering a sufficient concentration range were available for digitisation and subsequent calculation of an EC_90_ value. For the remainder it was not possible to calculate an EC_90_. Drugs with an available EC_90_ were ranked according to their Cmax/EC_90_ ratio (Figure 2). Drugs with a value above 1.0 achieved plasma concentrations above the concentrations reported to inhibit 90% of SARS-CoV-2 replication. Only eltrombopag, favipiravir, remdesevir, nelfinavir, niclosamide, nitazoxanide and tipranavir were estimated to exceed at least one of their reported EC_90_ by 2-fold or more at Cmax concentrations. Anidulafungin, lopinavir, chloroquine and ritonavir were also reported to exceed at least one of their reported EC_90_ values at Cmax but by less than 2-fold.

**Figure 2.**
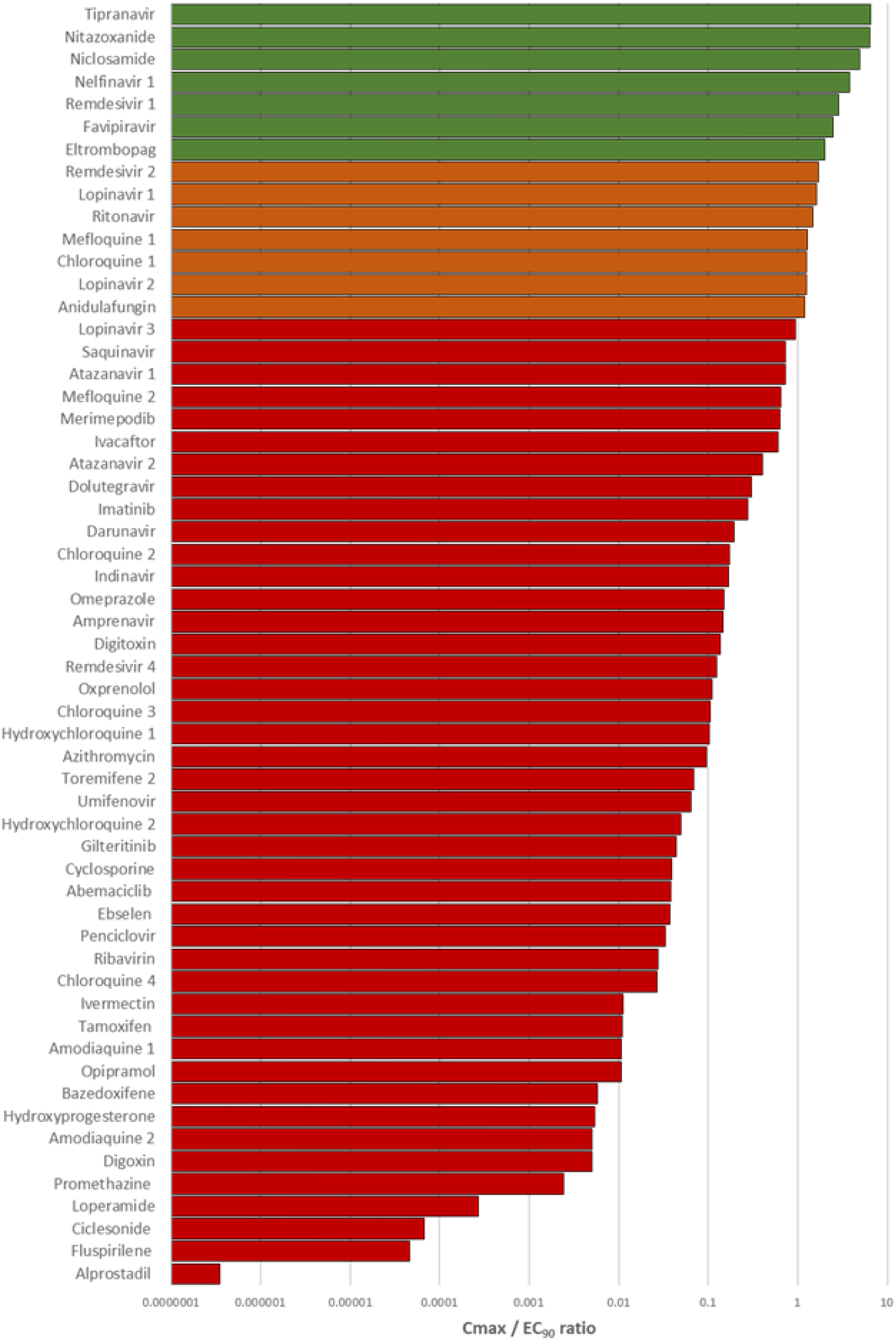
A bar chart displaying Cmax / EC_90_ ratio for compounds studied for *in vitro* antiviral activity against SARS-CoV-2 for which data were available to recalculate an EC_90_. Drugs with a ratio below 1 were deemed not to provide plasma concentrations at their approved doses to exert sufficient systemic antiviral activity. Those drugs with a ratio above 1 (shown in orange) were deemed to have potential to provide plasma concentrations sufficient to exert at least some antiviral activity for at least some of their dosing interval at their approved dose. Drugs shown in green were predicted to exceed plasma concentrations over their EC_90_ by more than a two-fold.

### Detailed interrogation of the plasma pharmacokinetics in relation to reported anti-SARS-CoV-2 activity

For drugs with Cmax concentrations above at least one of their reported EC_90_ values that are not already in clinical trials for Covid-19, a detailed evaluation of concentrations across their approved dosing interval was undertaken. For this, published pharmacokinetics data were digitised and replotted relative to the calculated EC_50_ and EC_90_ data for SARS-CoV-2 (Figure 3). For tipranavir (boosted with ritonavir), nelfinavir, sulfadoxine and nitazoxanide plasma concentrations after administration of the approved dose remained above SARS-CoV-2 effective concentrations across the entire dosing interval. For antifungdalin, etrombopag, lopinavir (boosted with ritonavir), mefloquine and chloroquine, Cmax values were above EC_90_ at 2, 6, 8, and 24 hours post dose respectively, but concentrations would be expected to dip below the EC_50_ at 3, 8, 10, 72 and 120 hours post dose respectively when given at approved doses and schedules. An overview of these drugs is presented in Figure 2.

**Figure 3.**
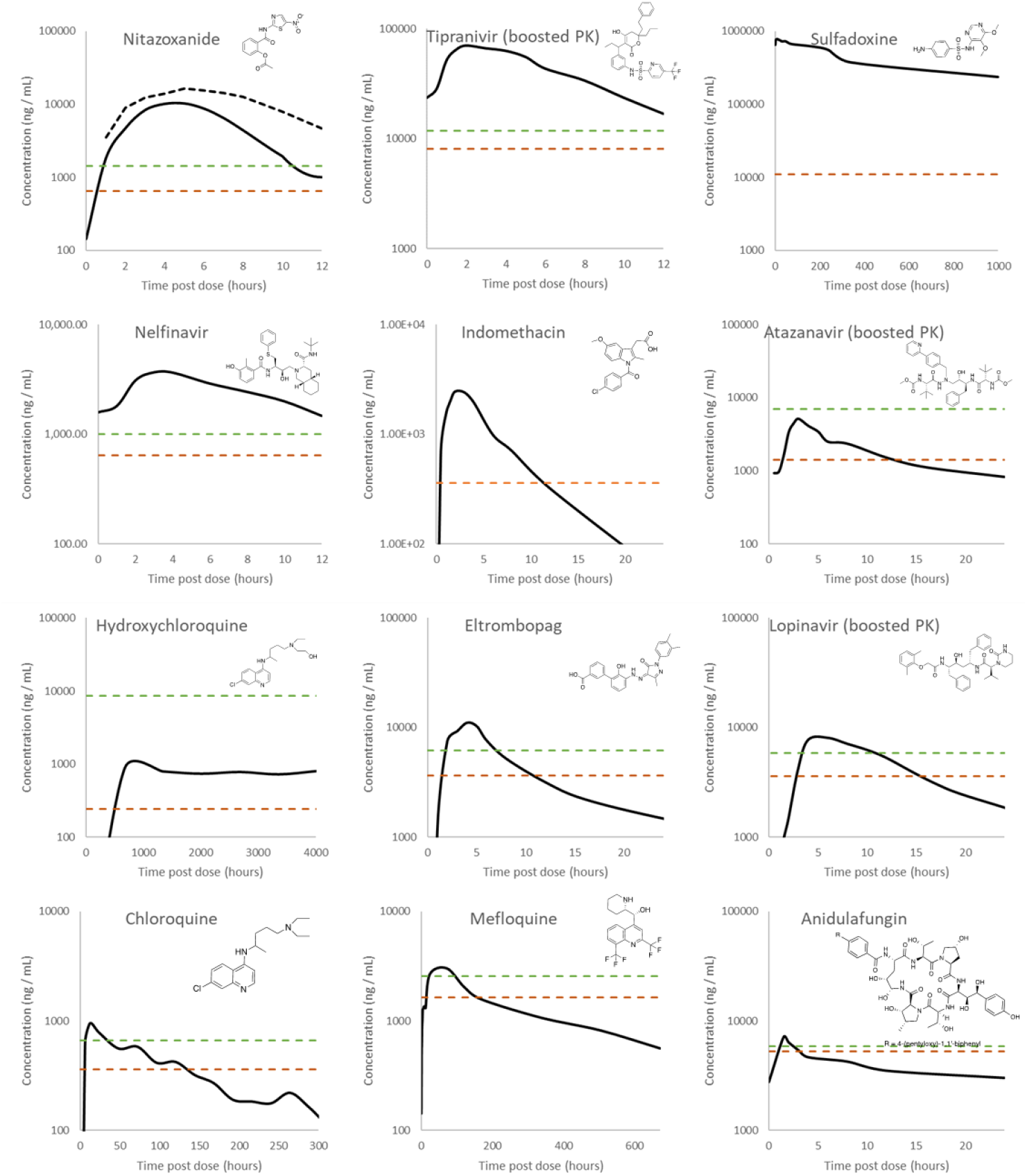
Digitised pharmacokinetic interrogation of all drugs calculated to have a Cmax / EC_50_ ratio above 1. The lowest reported SARS-CoV-2 EC_50_ (dashed orange lines) and associated recalculated EC_90_ (dashed green lines) are also highlighted. References for the utilised data are nitazoxanide 500mg BID and 1000mg BID [87], tipranavir 500mg BID with 200mg ritonavir [88], sulfadoxine 1500mg with 75mg pyrimethamine [72], nelfinavir 1250mg BID [89], indomethacin 50mg TID [90], atazanavir 300mg QD with 100mg ritonavir [91], hydroxychloroquine 2000 mg hydroxychloroquine sulfate / 1550 mg base administered over 3 days [92], Eltrombobag 75mg single dose [93], lopinavir 400 mg with 100 mg ritonavir [94], chloroquine 1500mg administered over 3 days [95], mefloquine 1200mg over 3 days [96], anidulafungin 100 mg QD [97]. Robust pharmacokinetic data were unavailable for niclosamide 500mg, ritonavir 600mg and merimepodib 300mg in order to conduct this digitised interrogation of these molecules.

### Simulated lung exposure relative to reported anti-SARS-CoV-2 activity

Lung KpU was simulated for all molecules for which the necessary physicochemical properties and *in vitro* drug binding information were available. KpU_lung_ was then used along with fraction unbound in plasma (Fu) and plasma Cmax values to calculate a predicted Cmax/EC_50_ (Figure 3) and Cmax/EC_90_ in lung (data not shown). For 4 drugs, ebselen, merimepodib, niclosamide and remdesivir, the fraction unbound data were unavailable. For 6 other drugs, benztropine, indinavir, loperamide, nelfinavir, saquinavir and toremifene the blood to plasma ratios were unavailable. For a further 4 drugs, camostat, emetine, fluspirilene and umifenovir both fraction unbound and blood to plasma ratios were unavailable. Therefore, these drugs were excluded from the analysis. A total of 23 drugs with available data were predicted to give concentrations in lung above at least one of their reported EC_50_ against SARS-CoV-2 (Figure 4) and 8 of these were predicted to exceed their EC_50_ by more than 10-fold. The rank order of lung Cmax/EC_90_ ratio was chloroquine > atazanavir (boosted with ritonavir) > tipranavir (boosted with ritonavir) > hydroxychloroquine > mefloquine > ivermectin > lopinavir (boosted with ritonavir) > azithromycin > ritonavir > gilteritinib > nitazoxanide > imantinib > oxprenolol (data excluded due to this analysis only being possible for 33 of the 56 drugs).

**Figure 4.**
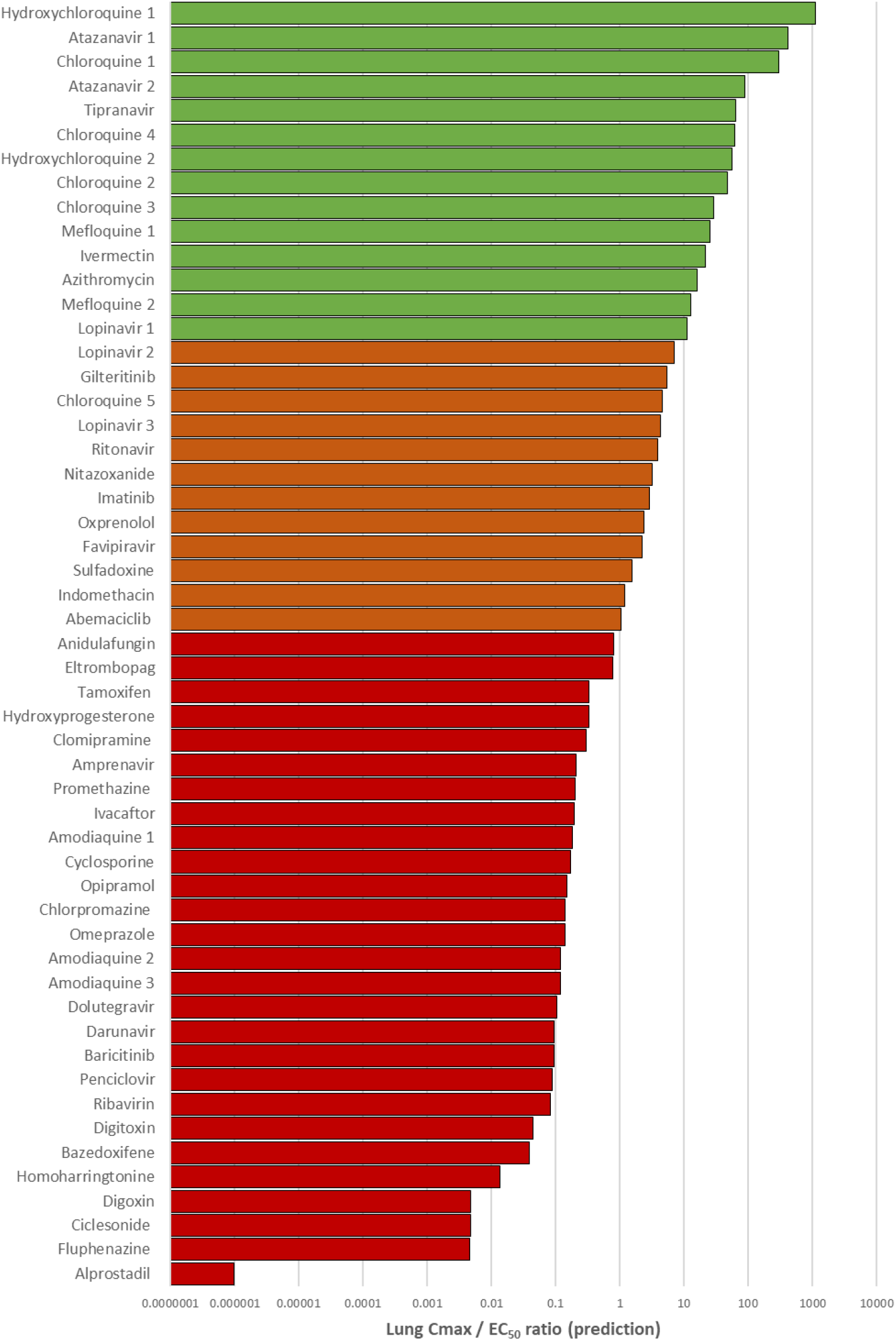
A bar chart displaying the simulated lung Cmax / EC_50_. Drugs with a ratio below 1 were deemed not to provide lung concentrations at their approved doses to exert sufficient pulmonary antiviral activity for treatment or prevention strategies. Those drugs with a ratio above 1 (shown in orange) were estimated to provide lung concentrations sufficient to exert at least some antiviral activity at their approved dose. Drugs shown in green were predicted to exceed lung concentrations over their EC_50_ by more than a ten-fold.

## Discussion

The systematic development of mechanism-based inhibitors for key targets involved in viral replication or pathogenesis is likely to result in highly effective and safe medicines in the coming years. However, the repurposing of already approved medicines in antiviral treatment or chemoprevention strategies is undoubtedly the fastest way to bring forward therapeutic options against the urgent unmet need posed by SARS-CoV-2. A range of different drugs and drug classes have been demonstrated to display varying degrees of antiviral activity against SARS-CoV-2 *in vitro*, and many of these drugs are already licenced for use in humans for a range of indications. However, currently the data emerging from global screening efforts are not being routinely benchmarked and prioritised against achievable concentrations after administration of doses proven to have acceptable safety profiles in humans.

The current analysis indicates that only 12 drugs with reported antiviral activity are likely to achieve plasma exposures above that required for antiviral activity for at least some of their dosing interval. Notably, neither chloroquine, hydroxychloroquine nor lopinavir/ritonavir exhibited a sustained plasma concentration above their reported SARS-CoV-2 EC_90_ across their reported dosing interval. Ultimately, the implications of this for therapy will depend upon whether systemic suppression is a prerequisite for a reduction in morbidity or mortality, but this does raise some concern for ongoing trials with these drugs (chloroquine: NCT04323527; NCT04333628 hydroxychloroquine: NCT04316377; NCT04333225; NCT04307693 and lopinavir/ritonavir: NCT04331834; NCT04255017; NCT04315948). However, the predicted lung accumulation rather than plasma exposure may provide some therapy advantage and/or give more reassurance for ongoing chemoprevention trials.

At least 5 of the 13 candidates achieving Cmax above one of their reported EC_50_ and derived EC_90_ are already in clinical evaluation for treatment of SARS-CoV-2. These include remdesivir (NCT04292730; NCT04292899; NCT04257656; NCT04252664; NCT04315948), favipiravir (NCT04310228; NCT04319900), lopinavir/ritonavir and chloroquine. No robust antiviral activity data were found for galidesrivir on which to conduct an analysis but it is also under clinical investigation (NCT03800173). A recent trial for favipirivir, demonstrated some success with an improvement over arbidol from 56% to 71% (p = 0.02) in patients without risk factors (but not critical cases or patients with hypertension and/or diabetes) [38]. The results of compassionate use of remdesivir in severely ill patients was also recently reported, and if confirmed in ongoing randomized, placebo-controlled trials will serve as a further validation of the other candidates presented here [39]. The authors are unaware of any ongoing COVID-19 trials for the other 7 candidate molecules with Cmax above their reported EC_90_, and none were reported on www.clinicaltrials.gov at the time of manuscript submission (14^th^ April 2020). Of particular interest nitazoxanide, tipranavir, sulfadoxine and nelfinavir may be expected to sustain their plasma pharmacokinetic exposure above their lowest reported EC_50_ and derived EC_90_ (where available) for the duration of their approved dose and dosing interval.

Nitazoxanide is an antiprotozoal drug that has previously been demonstrated to display broad antiviral activity against human and animal coronaviruses [40] as well as various strains of influenza [41, 42]. Importantly, nitazoxanide is rapidly metabolised to tizoxanide in humans and this active metabolite is being investigated against SARS-CoV-2 (NCT04341493 and NCT04343248). Tizoxanide has been reported to exhibit similar activities to nitazoxanide for other viruses as well as other pathogens [43-45]. The mechanism of antiviral action is not fully understood for nitazoxanide, but it has been reported to affect viral genome synthesis, prevent viral entry and interfere with the N-glycosylation and maturation of the influenza hemagglutinin [46-49]. Notably, the SARS-CoV-2 spike protein is also highly N-glycosylated [50]. This drug has also been shown to elicit an innate immune response that potentiates the production of type 1 interferons [46, 51] and a phase 2b/3 clinical trial demonstrated a reduction in symptoms and viral shedding in patients with uncomplicated influenza [42]. The safety of nitazoxanide is well understood, but it has not been fully investigated during renal or hepatic impairment. The antiviral activity of nitazoxanide for SARS-CoV-2 requires further study but the existing data for this drug are encouraging. Niclosamide is another antiprotozoal drug that exhibits broad antiviral activity due to its ability to perturb the pH-dependent membrane fusion required for virus entry [52] but it was reported to have no impact upon the attachment and entry of SARS-CoV-2 [53]. For MERS-CoV niclosamide was observed to inhibit SKP2 activity impairing viral replication [54]. Niclosamide has been reported to be well tolerated and does not influence vital organ functions [55]. However, it has low aqueous solubility and poor oral bioavailability [56] and despite a higher reported SARS-CoV-2 potency [28] than nitazoxanide [27], the Cmax/EC_90_ ratio was slightly lower. There is a paucity of published pharmacokinetic data for niclosamide and this prohibited a thorough investigation of exposures in relation to activity over its entire dosing interval. Both nitazoxanide and niclosamide have also been reported to be potent antagonists of TMEM16A, calcium activated chloride channels that modulate bronchodilation [57].

Tipranavir and nelfinavir are HIV protease inhibitors [58] and both drugs ranked highly in terms of their Cmax / EC_90_ ratio. Moreover, a more in-depth analysis demonstrated that the concentrations across the dosing interval for both these drugs remained above the calculated EC_90_ values at approved doses and schedules. Unlike nelfinavir, tipranavir has to be co-administered with a low dose of ritonavir to boost its pharmacokinetics via CYP3A4 inhibition [59]. Since ritonavir itself has been reported to exert anti-SARS-CoV-2 activity, this could be advantageous, but would need to be balanced against the much higher risk of drug-drug interactions that could negatively impact patient management. The implications of drug interactions have already been raised for this reason with lopinavir/ritonavir use for COVID-19 [60] and are likely to be exacerbated with the higher ritonavir dose needed for tipranavir. Moreover, tipranavir has a black box warning from the FDA for fatal and nonfatal intracranial haemorrhage as well as severe hepatotoxicity [61-63]. The major route of metabolic clearance for nelfinavir is via CYP2C19 and this pathway generates the M8 metabolite that retains activity against the HIV protease [64]. No data are available for inhibition of SARS-CoV-2 replication by the M8 metabolite but if active, this could provide and advantage for nelfinavir over tipranavir for COVID-19. Conversely, While the analysis of pharmacokinetics relative to potency of these molecules against SARS-CoV-2 is encouraging, it should be noted that the reported *in vitro* activity for HIV [58, 65] is far higher than that against SARS-CoV-2 and both drugs are highly protein bound [66, 67]. While older HIV protease inhibitors like tipranavir and nelfinavir are associated with long-term toxicities [58, 68-70] these may be less of a concern over shorter term exposure if they prove to be successful for COVID-19.

Sulfadoxine is another antimalarial drug that is usually administered in combination with pyrimethamine as a folic acid antagonist combination [71]. Sulfadoxine inhibits the activity of dihydropteroate synthase within the malaria parasite, but its mechanism of action for SARS-CoV-2 is unclear. It should also be noted that the authors can find no data describing antiviral activity of this drug against other viruses. Also, the concentrations used in the *in vitro* activity used in this analysis [26] were not high enough to reach or calculate an EC_90_ value. Therefore, like other molecules described in this manuscript *in vitro* anti-SARS-CoV-2 activity should be repeated. Notwithstanding, sulfadoxine plasma concentrations far above the reported EC_50_ are maintained in patients receiving a single 1500 mg dose (with 75 mg pyrimethamine) for over 40 days [72]. Compared to some other reported molecules sulfadoxine is not expected to have as high an accumulation in the lungs, but concentrations higher than its EC_50_ are estimated from the analysis of its lung KpU. Therefore, if the reported antiviral activity is confirmed this drug may offer opportunities for therapy and/or chemoprophylaxis.

Considering that most of the impact of severe disease occurs in the lung and that this tissue may be a key site for transmission, the potential of candidate drugs to accumulate in lung tissue was considered. The analysis of predicted lung Cmax / EC_50_ ratio revealed more candidates expected to exceed the concentrations needed for antiviral activity in this tissue. Hydroxychloroquine, chloroquine, mefloquine, atazanavir (boosted with ritonavir), tipranavir (boosted with ritonavir), ivermectin and lopinavir were all predicted to achieve lung concentrations over 10-fold higher than their reported EC_50_. All of these drugs were also predicted to exceed their EC_90_ in lung by at least 3.4-fold (data not shown). The lung prediction was not possible for nelfinavir because insufficient data were available to calculate K_p_U_lung_ but nitozoxanide and sulfadoxine were also predicted to exceed their reported EC_50_ by 3.1- and 1.5-fold in lung, respectively. Nitazoxanide was predicted to exceed its EC_90_ by 1.4-fold in lung but an EC_90_ was not calculable from the available data for sulfadoxine.

During inflammation or injury, changes to the vascular microenvironment, could have a profound effect on the ability of these drugs to accumulate in lung cells. Due to the recruitment of neutrophils and leaky endothelial cells [73], the lung inflammatory microenvironment is characterized by increased body temperature, excessive enzymic activity and most importantly, by a low interstitial pH [74]. In the case of chloroquine and hydroxychloroquine, these diprotic weak bases are exquisitely dependent on a pH gradient to drive lysosomal uptake as a mechanism of lung accumulation. It has been demonstrated that cellular chloroquine uptake is diminished one hundred-fold for every pH unit of external acidification [75]. This situation is likely to deteriorate further on mechanical ventilation, which also induces acidification of the lung tissue, independently of inflammation [76, 77]. Therefore, the benefits of lung accumulation for many of these drugs may be lost during treatment of severe SARS-CoV-2 infection. Conversely, mefloquine is monoprotic and more lipophilic than chloroquine, which may make it much less reliant on the pH gradient to drive cellular accumulation in lung. It is likely that the charged form of the drug is sufficiently lipophilic to allow movement across biological membranes along a concentration gradient [78]. Only two studies have described mefloquine uptake into cells, one study suggested that mefloquine uptake is not energy-dependent and the other suggested that mefloquine uptake is mediated by secondary active transport, rather than passive proton trapping [79, 80]. Mefloquine is known to cause severe psychiatric side effects in some patients and so use of this drug should be managed with care [81]. Therefore, mefloquine may offer opportunities for treatment during severe disease that are not available with other drugs currently being tested for COVID-19 therapy. If the high lung exposures are proven empirically for the drugs on this list, then some may also prove to be valuable for chemoprevention strategies.

### Limitations of this analysis

This study represents the first holistic view of drugs with reported activity against SARS-CoV-2 in the context of their achievable pharmacokinetic exposure in humans. While the analysis does provide a basis to rationally selected candidates for further analysis, there are some important limitations. Firstly, Cmax was the only pharmacokinetic parameter that was universally available for all of the candidate drugs, but Cmin values are generally accepted as a better marker of efficacy since they represent the lowest plasma concentration over the dosing interval. However, Cmax was only used to assess whether plasma concentration would exceed those required at any point in the dosing interval, and this was followed by a more in-depth analysis of the most promising candidates.

Secondly, an EC_50_ value only equates to a concentration required to suppress 50% of the virus, and data were unavailable to calculate EC_90_ values for some of the drugs. EC_90_ values are a preferred marker of activity because the slope of the concentration-response curve can vary substantially between different molecules and between different mechanisms of action. Although EC_90_ were not calculable for all drugs the authors deemed it appropriate to deprioritise molecules not achieving EC_50_ at Cmax in this analysis. Thirdly, the reported antiviral activities were conducted under different conditions (Table 1) and in several cases varied between the same molecule assessed in different studies (Figure 1). No mitigation strategy was possible for this limitation and the data should be interpreted in the context that the quality of the available data may profoundly impact the conclusions. Furthermore *in vitro* activity should be confirmed for the promising candidates.

Fourthly, plasma protein binding can be an important factor in determining whether sufficient free drug concentration are available to exert antiviral activity [86] and insufficient data were available across the dataset to determine protein binding-adjusted EC_90_ values. This is important because for highly protein bound drugs the antiviral activity in plasma may be lower than reported *in vitro* activity because protein concentrations used in culture media are lower than those in plasma. Fifthly, robust pharmacokinetic data were not available for all the molecules and subtle differences have been reported in the pharmacokinetics in different studies. Where possible, this analysis utilised the pharmacokinetics described at the highest doses approved for other indications and checked them to ensure that profound differences were not evident between different studies. However, in some cases higher doses and/or more frequent dosing has been investigated for some of the drugs mentioned so higher exposures may be available for some drugs with off-label dosing. Sixthly, the digitised pharmacokinetic plots presented in this manuscript represent the mean or median profiles depending on what was presented in the original manuscripts. Many of the drugs presented are known to exhibit high inter-individual variability that is not captured within the presented analysis and it is possible that even for promising candidates a significant proportion of patients may have sub-therapeutic concentrations despite population mean/median being higher than the Cmax. Advanced pharmacokinetics modelling approaches will be needed to unpick this and are currently underway by the authors.

Seventhly, the presented predictions for lung accumulation may offer a basis for ranking molecules for expected accumulation in that organ, but ultimate effectiveness of a chemoprophylactic approach will likely depend upon penetration into other critical matrices in the upper airways, for which there are currently no robustly validated methods of prediction. Also, while a generally accepted method for assessing KpU was employed, the predictions are not supported by any empirical investigation of the lung accumulation.

Finally, this analysis assumes that drugs need to be active within the systemic compartment in order to have efficacy against SARS-CoV-2. Since current evidence suggests that the virus is widely disseminated throughout the body this is a logical assumption. However, ultimate efficacy of any drug can only be demonstrated with robust clinical trial designs.

## Summary

The current analysis reveals that many putative agents are never likely to achieve target concentrations necessary to adequately suppress SARS-CoV-2 under normal dosing conditions. It is critical that candidate medicines emerging from *in vitro* antiviral screening programmes are considered in the context of their expected exposure in humans where possible. Clinical trials are extremely time consuming and expensive, and it is critical that only the best options are progressed for robust analysis as potential mono- or combination therapy or prevention options. Finally, it would be highly beneficial for activity data for SARS-CoV-2 to be performed with standardised protocol and with activity reported as EC_90_ values as a better marker of the concentrations required to suppress the virus to therapeutically relevant levels.

## Data Availability

The data that support the findings of this study are available from the corresponding author upon reasonable request.

